# Circulating CD8+ MAIT cells correlate with improved outcomes in anti-PD1 treated melanoma patients

**DOI:** 10.1101/2020.08.20.20178988

**Authors:** Victoria M. Vorwald, Dana M. Davis, Robert J. Van Gulick, Robert J. Torphy, Jessica S.W. Borgers, Jared Klarquist, Kasey L. Couts, Carol M. Amato, Dasha T. Cogswell, Mayumi Fujita, Timothy Davis, Catherine Lozupone, Theresa M. Medina, William A. Robinson, Laurent Gapin, Martin D. McCarter, Richard P. Tobin

## Abstract

While much of the research concerning factors associated with responses to immunotherapies focuses on the contributions of conventional peptide-specific T cells, the role of unconventional T cells, such as mucosal-associated invariant T (MAIT) cells, in human melanoma remains largely unknown. MAIT cells are innate-like T cells expressing a semi-invariant T cell receptor restricted to the non-classical MHC class I molecule MR1 presenting vitamin metabolites derived from bacteria. In this prospective clinical study, we sought to characterize MAIT cells in melanoma patients and determine their association with clinical outcomes. We identified tumor-infiltrating MAIT cells in melanomas across metastatic sites and found that the number of circulating MAIT cells is reduced in melanoma patients. However, circulating MAIT cell frequency is restored by anti-PD1 treatment in responding patients, correlating with treatment responses in which patients with high frequencies of MAIT cells exhibited improved overall survival. These data provide evidence for leveraging MAIT cells and their functions as novel targets for future therapies.

## Introduction

While recent efforts to identify characteristics of melanoma patients that are associated with differential clinical responses to immune checkpoint inhibitors have led to significant clinical advances, further analysis is required to elucidate the complexity of immune reactions that result in successful antitumor immunity. One newly described population of cells that is highly abundant in humans and yet missing from previous analyses is mucosal-associated invariant T (MAIT) cells. Unlike conventional T cells that express highly variable T cell antigen receptors (TCRs) that recognize peptide antigens presented by highly polymorphic conventional HLA class I and II molecules, MAIT cells are a population of innate-like T cells with a restricted TCR repertoire (Gapin, 2014; Godfrey et al., 2019; Kjer-Nielsen et al., 2012; Reantragoon et al., 2013; Reantragoon et al., 2012), that recognize small metabolite molecules derived from bacterial riboflavin synthesis presented by the non-classical and non-polymorphic MHCI-related molecule MR1(Godfrey et al., 2019; Kjer-Nielsen et al., 2012; Krovi and Gapin, 2016; Reantragoon et al., 2012; Tastan et al., 2018). MAIT cells comprise up to 10% of peripheral blood CD8+ T cells in healthy adults and accumulate at mucosal sites including the gut, lungs, and liver, where they can account for up to 50% of CD8+ T cells (Kurioka et al., 2016). This high frequency makes MAIT cells the most abundant currently known population of T cells with a single antigen specificity (Koay et al., 2018). In humans, MAIT cells are accurately identified by staining with MR1-tetramers loaded with the microbial ligand 5-(2-oxopropylideneamino)-6-D-ribitylaminouracil (5-OP-RU) (Corbett et al., 2014). Upon recognition of their cognate ligand, MAIT cells rapidly respond by producing cytokines including tumor necrosis factor-α (TNFα), interferon-γ (IFNγ) and interleukin-17 (IL)-17, and upregulate CD40 ligand (CD40L, CD154) (Godfrey et al., 2019; Kurioka et al., 2016), suggesting that they can also indirectly affect the activation status and function of other immune cells (Salio et al., 2017). MAIT cells can also have direct cytotoxic functions on both virally-infected and tumor cells (Rudak et al., 2018). Finally, due to their primed/memory nature, MAIT cells express high levels of several cytokine receptors, including IL-18Rα, IL-12R and IL-15R, which, upon engagement, can lead to MAIT cell activation in the absence of TCR triggering, resulting in the rapid production of IFNγ, TNFα, and IL-17 and cytotoxic functions including the production of perforin, granzyme B (GZMB), and granzyme K (GZMK) (Godfrey et al., 2019; Sattler et al., 2015). Additionally, MAIT cells have been shown to activate antigen-presenting cells capable of promoting antitumor immunity (Salio et al., 2017).

Previous studies of MAIT cells in human cancer patients and mouse models of cancer have revealed mixed results (Gherardin et al., 2018; Melo et al., 2019; Yan et al., 2020). In a mouse model of melanoma, MAIT cells were found to exert tumor-promoting functions, with slower progressing tumors and fewer metastases in MAIT cell-deficient MR1-/- mice (Yan et al., 2020). Similarly, hepatocellular carcinoma patients with high levels of tumor-infiltrating MAIT cells had worse overall survival compared to those with low infiltrating MAIT cell levels (Duan et al., 2019). However, an opposing result was found in patients diagnosed with esophageal cancer, multiple myeloma, or colorectal cancer, where higher levels of MAIT cells were associated with decreased disease burden and improved outcomes (Gherardin et al., 2018; Ling et al., 2016; Melo et al., 2019). Further confounding this topic, a recent study in colorectal cancer patients found that a regulatory subset of CD4+ MAIT cells is activated within the tumor microenvironment and the abundance of these cells is correlated with tumor bacterial load (Li et al., 2020). To date, the role of MAIT cells in responses to immune checkpoint inhibitors (ICIs) in human cancer patients is unknown.

In this prospective study, we sought to determine the frequency and functional status of MAIT cells in patients with melanoma, and whether it might correlate with clinical outcomes. We also investigated the activation of MAIT cells in the peripheral blood and tumors from a variety of metastatic sites in melanoma patients. We report that MAIT cells are reduced in the circulation of melanoma patients. However, circulating MAIT cell levels were restored in patients responding to ICI therapy and higher frequencies of circulating MAIT cells were associated with improved overall survival. Altogether, our results reveal circulating MAIT cell frequencies as a potential prognostic correlate of therapeutic responses to anti-PD1 immunotherapy and suggest that manipulating MAIT cells might be advantageous to anti-melanoma immune responses.

## Results and discussion

### The frequency of MAIT cells is decreased in the circulation of melanoma patients

MAIT cells were identified using flow cytometry as live, CD45^+^CD3^+^CD161^+^5-OP-RU-Tetramer^+^ cells, and further stratified into subsets based on CD4 and CD8 expression (Fig. 1a). The frequency of MAIT cell subsets was analyzed in the peripheral blood of healthy donors (HD, n = 11) and treatment naïve (TN, n = 33) melanoma patients regardless of stage. We found that the frequency of circulating total (Fig. 1b) and CD8+ MAIT cells (Fig. 1d) was reduced in melanoma patients compared to HD. The reduction in these populations was apparent regardless of whether it was expressed as a proportion of the total CD3^+^ T cells or of total CD45^+^ cells. We observed no statistically significant changes in the frequency of CD4+ MAIT cells as proportions of CD4+ T cells, total T cells, or CD45^+^ cells (Fig. 1c). However, the frequency of CD4+ MAIT cells as a proportion of MAIT cells was increased in melanoma patients compared to HD (Fig. 1e), though this did not reach statistical significance (*p* = 0.069). Conventional (5-OP-RU-MR1 Tetramer^−^) non-MAIT (NM)-CD8+ and NM-CD4+ T cells are usually associated with the capacity to mount antitumor immune responses (Lu et al., 2014; Sharma and Allison, 2020; Wei et al., 2019). To determine if the observed frequency changes were specific to MAIT cells or reflected a more general T cell lymphopenia in melanoma patients, we compared the frequency of NM-CD8+ and NM-CD4+ T cells in HD and melanoma patients. No significant differences in the frequency of either NM-CD4+ or NM-CD8+ T cells was observed between HD and melanoma patients (Supplementary Fig. S1a and S1b), demonstrating that the decreased frequency of MAIT cells in the blood of melanoma patients is specific to this T cell population.

**Figure 1.**
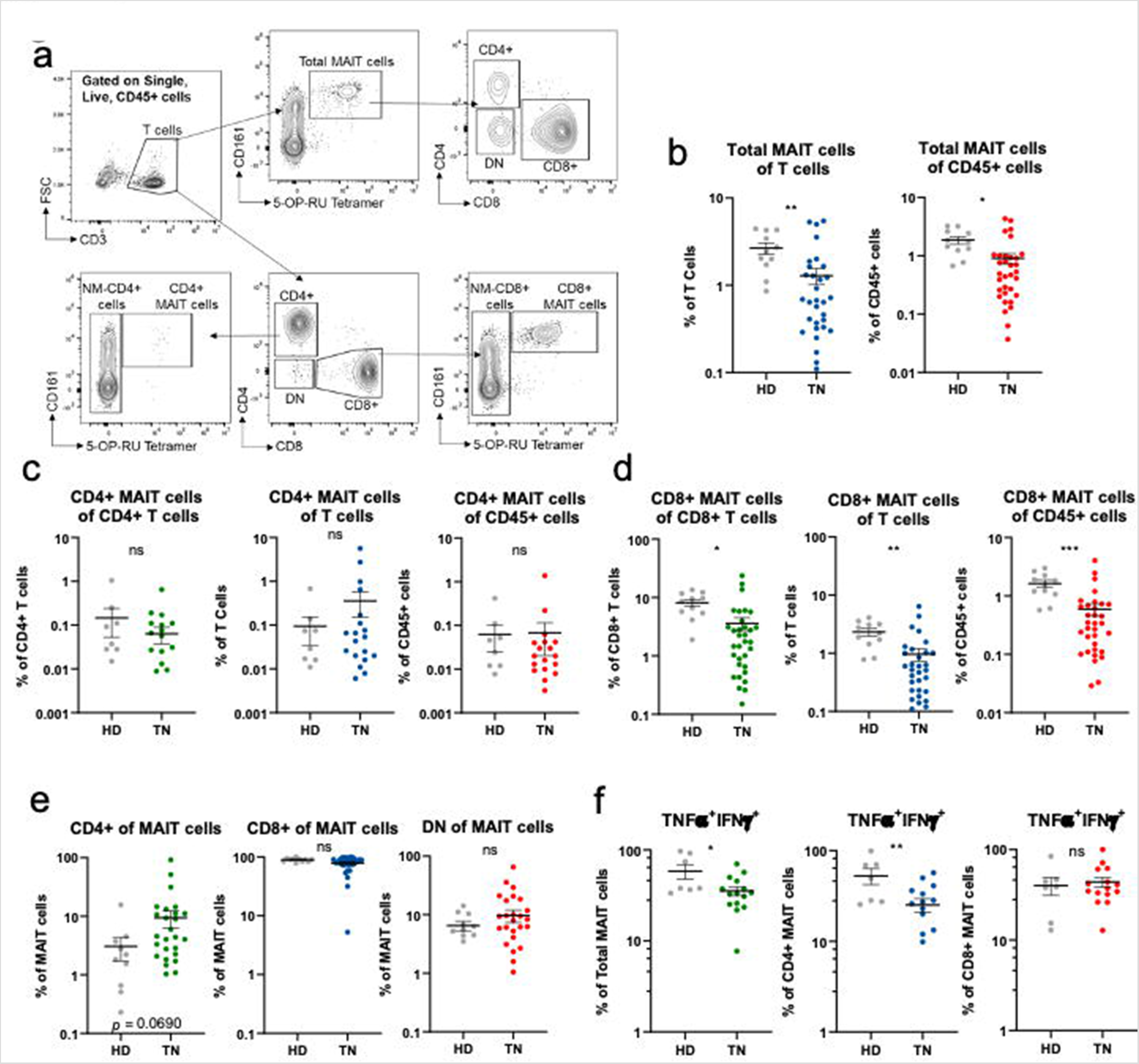
MAIT cells are decreased in the circulation of melanoma patients. (**a**) Example flow cytometric gating strategy to identify MAIT cell subsets in peripheral blood. Comparisons of Total (**b**), CD4+ (**c**), and CD8+ (**d**) MAIT cell populations in healthy donors (HD) and treatment naïve (TN) melanoma patients as a percentage of the indicated population. (**e**) Comparison of the ratio of CD4+, CD8+, and double negative (CD4^−^CD8^−^, DN) among MAIT cells. (**f**) Comparisons of the frequency of polyfunctional (TNFα+INFγ+) cells as a percentage of the described population.

### Melanoma-dependent changes in MAIT cell frequencies are associated with decreased cytokine production

Human MAIT cells have been reported to produce the cytokines IFNγ, TNFα, and IL-17, as well as the effector molecules GZMB and GZMK upon activation (Bulitta et al., 2018; Godfrey et al., 2019; Sundstrom et al., 2015). To determine if there were functional differences in MAIT cells in melanoma patients, we measured the expression of IFNγ, TNFα, GZMB, and GZMK in MAIT cells from healthy donors and melanoma patients. The percentages of polyfunctional total MAIT cells and CD4+ MAIT cells producing IFNγ and TNFα were reduced in melanoma patients regardless of stage compared to HD, while no changes in the production of these cytokines by CD8+ MAIT cells was observed (Fig. 1f). Similarly, no difference in the production of GZMB or GZMK in either circulating MAIT cells or NM-CD8+ T cells between HD and melanoma patients was observed regardless of stage (Supplementary Fig. S1c and S1d). These data suggest that while certain MAIT cell populations from melanoma patients have decreased cytokine production, cytotoxic capacity is maintained.

### MAIT cells infiltrate melanoma tumors

Melanoma metastasis can occur in nearly all tissues in the body (Damsky et al., 2010). While MAIT cells could be detected within melanoma metastases, in general their frequencies out of total intra-tumoral CD45^+^ cells were lower compared to what was observed in the circulation, and this effect was particularly marked for CD8+ MAIT cells (Fig. 2a and 2b). Instead, a trend towards an increased ratio of CD4+ MAIT cells within tumors was observed (Fig. 2a), with CD4+ and double negative (CD4^−^CD8^−^, DN) MAIT cell subsets constituting a significantly greater percentage of the tumor-infiltrating MAIT cells than in the circulation (Fig. 2c and 2d). These results resonate with recent findings in colon cancer where MAIT cells, and in particular regulatory CD4 MAIT cells, were also found preferentially in the tumor tissue (Li et al., 2020).

**Figure 2.**
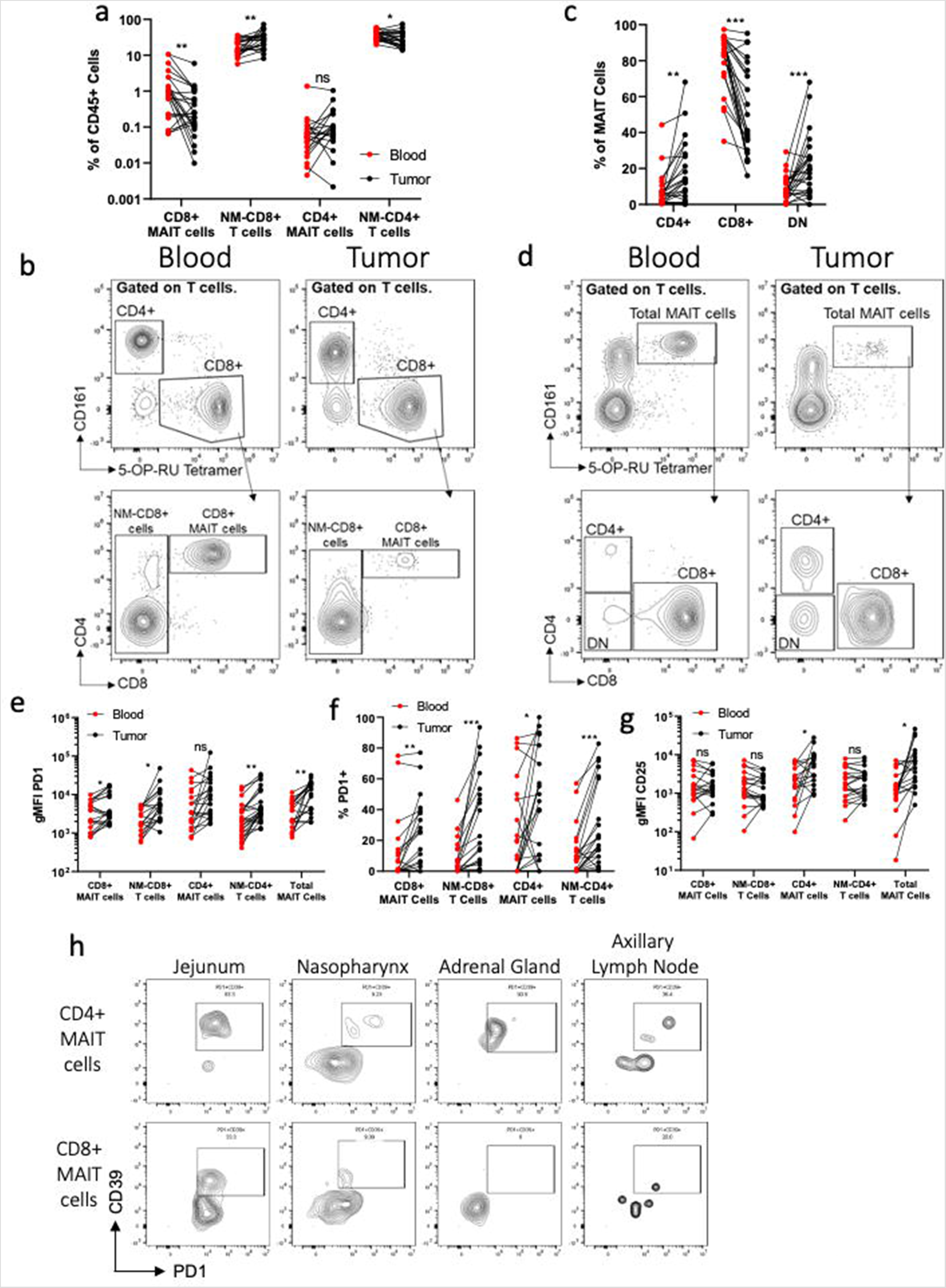
Activated MAIT cell infiltrate human melanoma tumors. (**a**) The frequency of MAIT cell subsets and their non-MAIT (NM) counterparts as a percentage of the total CD45+ cells in the blood and infiltrating the tumors. (**b**) Example staining of MAIT cells comparing blood vs. tumor. (c) The ratio of each MAIT cell subset as a percentage of MAIT cells comparing blood vs. tumor. (d) Example staining of MAIT cells comparing blood vs. tumor. Comparisons of the geometric mean fluorescence intensity (gMFI) (e) of PD1 on MAIT cell subsets or percentage of PD1+ cells in the blood vs. tumor. (g) The gMFI of CD25 on MAIT cell subsets in the blood vs. tumor. (h) Example staining of CD39+PD1+ cells in different metastatic sites gated on CD4+ or CD8+ MAIT cells.

### MAIT cells express higher levels of PD1 within the tumor microenvironment

We next characterized the phenotypic differences between MAIT cells in the blood and those infiltrating melanoma metastases. Total and CD8+ MAIT cells as well as NM-CD4+ and NM-CD8+ T cells, but not CD4+ MAIT cells, showed increased levels of PD1 expression within the tumor microenvironment compared to circulating cells (Fig. 2e). Interestingly, only CD4+ MAIT cells demonstrated increased CD25 expression, while CD154 levels were comparable on all T cell populations in the circulation and the tumor microenvironment (Fig. 2g and Supplementary Fig. S1e). Increased PD1 expression and low levels of CD25 and CD154 on conventional CD8+ T cells has been associated with chronic TCR stimulation in an immunosuppressive microenvironment (Ahn et al., 2018; Khan et al., 2019).

Therefore, the phenotype of tumor-infiltrating MAIT cells could also reflect chronic TCR stimulation, although we cannot exclude that other mechanisms, such as the tumor microenvironment or currently unknown factors may also be responsible. It is important to note that melanoma metastases were surgically resected as part of standard of care treatment and were generally removed when the lesion was not responding to anti-PD1 immunotherapy.

MR1-restricted T cells using different TCRs from the ones expressed by MAIT cells were recently shown to recognize tumor cells in an MR1-dependent manner (Crowther et al., 2020), suggesting that tumor-derived antigenic metabolites that can be presented by MR1 do exist. Whether such tumor-derived compounds also contain potential MAIT antigens is currently unknown. A recent report from the Newell group showed that the frequency of MAIT cells co-expressing CD39 and PD1 correlates with intratumoral bacterial load in colon cancer (Li et al., 2020). While we observed higher frequencies in melanoma tumors from mucosal sites (nasopharynx and jejunum), CD4+, and to a lesser extent CD8+, CD39^+^PD1^+^ MAIT cells were also observed in metastases to the adrenal gland and axillary lymph node, which are not normally associated with bacterial colonization (Fig. 2h). In their report, Li *et al*. proposed that bacterial ligand-dependent TCR stimulation drove CD39 expression on MAIT cells. The presence of CD39^+^PD1^+^MAIT cells in our samples could also be suggestive of TCR recognition of an unknown tumor-derived antigen. However, it cannot be excluded that such TCR stimulation might be occurring elsewhere in the body with subsequent circulation of activated CD39^+^PD1^+^MAIT cells through these sites, or that bacteria or bacteria-derived antigens are present within these metastatic sites. Furthermore, CD39 expression has been shown to be associated with T cell exhaustion (Canale et al., 2018), and may indicate an exhausted MAIT cell phenotype, similarly to what has been observed in other tumor types (Duan et al., 2019). Nevertheless, MAIT cells can also be activated in the absences of TCR-stimulation via IL-18 and IL-12, as well as in combination with IL-15, resulting in the rapid production of IFNγ, TNFα, and IL-17 and cytotoxic (GZMB, GZMK) functions (Godfrey et al., 2019; Sattler et al., 2015).

### Metastatic melanoma patients responding to anti-PD1 immunotherapy have increased frequencies of circulating CD8+ MAIT cells

We next sought to determine if the decreased frequency of MAIT cells observed in the circulation of melanoma patients could be influenced by anti-PD1 immunotherapy and whether this might correlate with responses to treatment. Although it did not reach statistical significance in our current analysis, we found a trend towards an increased frequency of total MAIT cells (*p* = 0.061) in patients who went on to respond (R, n = 9), but not in non-responding patients (NR, n = 6), following the third anti-PD1 infusion compared to pre-treatment (Fig. 3a and 3b). However, closer examination of MAIT cell subsets revealed a statistically significant increase in the frequency of circulating CD8+ MAIT cells in responding patients but not in non-responders, with a reciprocal increased frequency of CD4+ MAIT cells in non-responders but not in responders (Fig. 3a and 3b). This result corroborates data from *in vitro* studies using MAIT cells isolated from multiple myeloma patients showing increased MAIT cell proliferation in the presences of anti-PD1 (Favreau et al., 2017). While the functional differences (i.e. cytokine production, cytotoxic functions, and tumor infiltration capacity) between CD4+ and CD8+ MAIT cells have not been fully explored in cancer patients, this is an active area of ongoing clinical and translational research.

**Figure 3.**
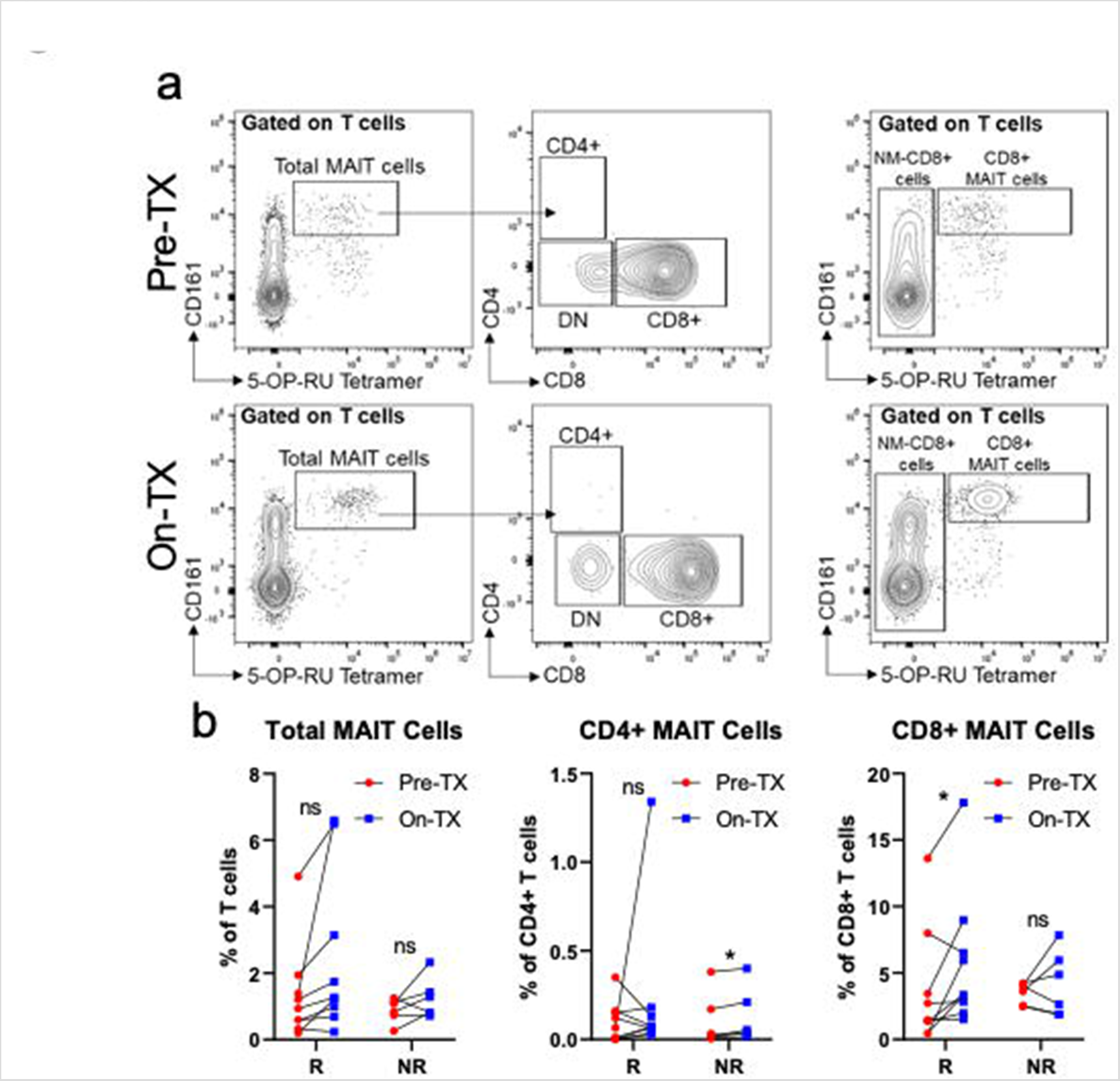
MAIT cells increase in frequency in the circulation of melanoma patients during anti-PD1 therapy. (a) Example flow cytometric data characterizing the frequency of circulating MAIT cell subsets prior to treatment (Pre-TX) and after the third anti-PD1 treatment (On-TX) in stage IV melanoma patients. (b) Comparisons of the labelled circulating MAIT cell subsets comparing Pre-TX and On-TX samples between responding (R) and non-responding patients. Response to anti-PD1 was characterized using RECIST1.1 criteria with responders identified as complete response (CR) or partial response (PR), while non-responders were identified as stable disease (SD) or progressive disease (PD).

### Melanoma patients responding to immune checkpoint inhibitors have increased frequencies and absolute numbers of circulating Total and CD8+ MAIT cells

To confirm these results from our smaller pre- and on-treatment blood draw cohort, and because we observed a strong reduction in the frequency of MAIT cells in melanoma patients, we next sought to determine if the circulating frequency of MAIT cell subsets was altered in early stage disease (stage I/II), advanced disease (stage III/IV), or in patients who respond to anti-PD1 therapies compared to those who do not. We found that total MAIT cells, expressed as either a proportion of T cells or as the absolute number of these cells in the circulation, are increased in patients who respond (n = 23) to anti-PD1 therapies compared to non-responders (n = 21) and treatment naïve patients (n = 17) (Fig. 4a and 4b). While the frequencies and absolute numbers of NM-CD8+ T cells, NM-CD4+ T cells, and either DN or CD4+ MAIT cells were not significantly different between responders and non-responders (Fig. 4a, 4c, and Supplementary Fig. S1f and S1g), we found that CD8+ MAIT cells, expressed as a proportion CD8+ T cells or total T cells, are significantly increased in responders compared to non-responders (Fig. 4a and 4d). Additionally, in responders, the increased frequency of total and CD8+ MAIT cells mirrored the increased absolute number of these cells in the circulation compared to treatment naïve or non-responding patients (Fig. 4a, 4b, and 4d).

**Figure 4.**
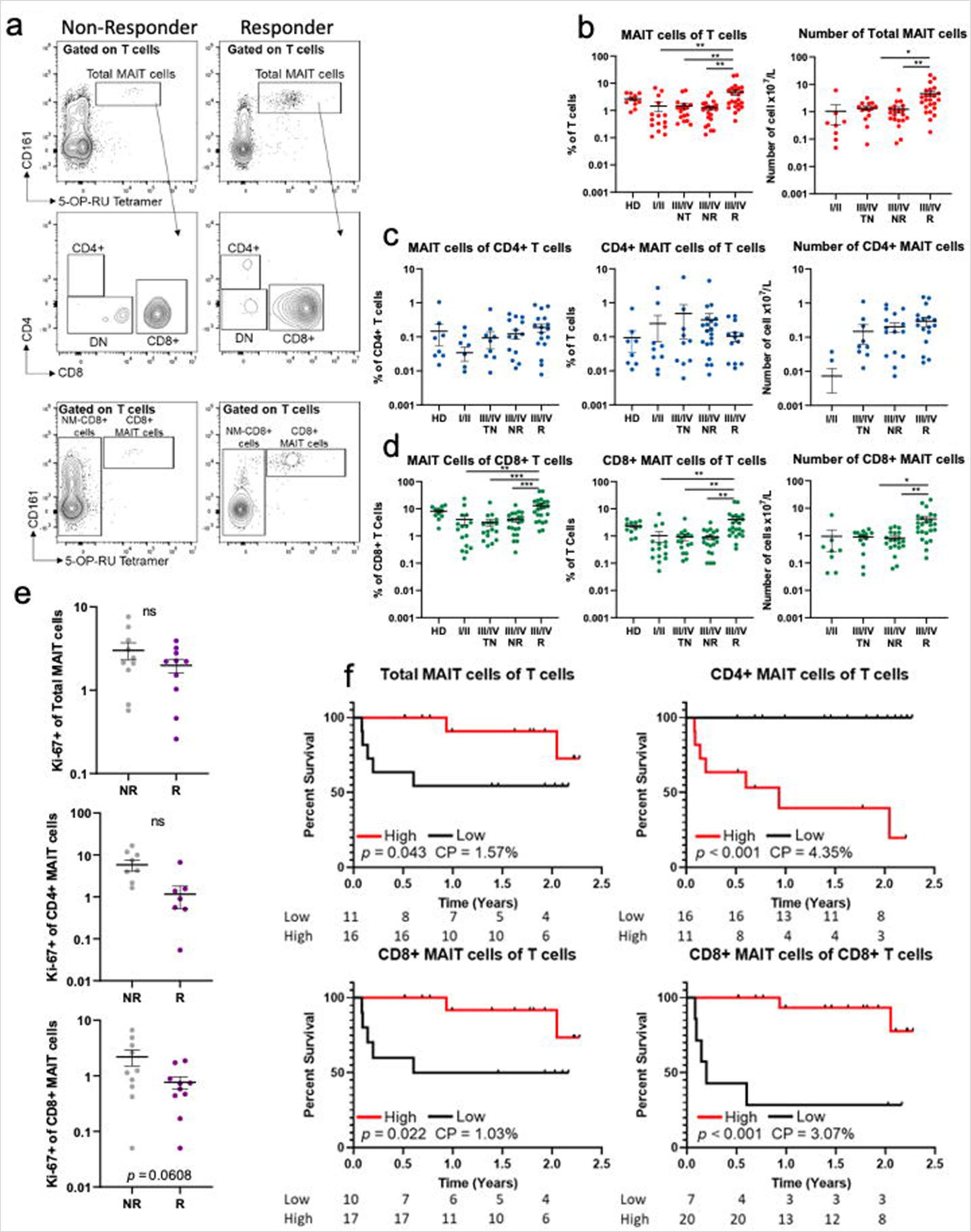
The frequency of Total and CD8+ MAIT cells is positively associated with improved clinical responses and overall survival in melanoma patients. (a) Example staining of circulating MAIT cell subsets in responding vs. non-responding melanoma patients. Comparisons of Total (b), CD4+ (c), and CD8+ (d) MAIT cell populations in healthy donors (HD), early stage (I/II), treatment naïve advanced stage (III/IV TN), advanced stage disease anti-PD1 non-responders (III/IV NR), and advanced stage disease anti-PD1 responders (III/IV R) as a percentage of the indicated population or the absolute number of each subset. (e) Comparisons of the frequency of proliferating cells (Ki-67+) in anti-PD1 non-responders (NR) compared to responders (R). (f) Kaplan-Meier curves comparing the survival of stage IV melanoma patients who received at least three doses of anti-PD1 therapy based on the frequency of the listed MAIT cell population. The number of patients at risk at each timepoint is indicated below each graph. The cutpoint (CP) for each cellular population is indicated on each graph. Response to anti-PD1 was characterized using RECIST1.1 criteria with responders identified as complete response (CR) or partial response (PR), while non-responders were identified as stable disease (SD) or progressive disease (PD).

Surprisingly, the increased frequency and total cell number of MAIT cells in the circulation of melanoma patients that are responding to anti-PD1 immunotherapy was not due to increased proliferation of the MAIT cells, as measured by Ki-67 staining (Fig. 4e). Instead, we found a trend towards and a proportion of Ki-67^+^ MAIT cells in the non-responding patients (Fig. 4e). There are at least two possible explanations for this observation. First, there is a possibility that MAIT cells in the responding patients are leaving the tissues and are reentering the circulation. Second, it is also possible that MAIT cells in the responders were less susceptible to cell death and therefore do not need to proliferate to maintain their normal levels. Previous work in HIV+ individuals has shown that MAIT cells are susceptible to PD1-induced cell death (Saeidi et al., 2015), and it is possible that blocking PD1:PD-L1 interactions prevents this from happening. However, both mechanisms remain to be fully elucidated.

### Circulating CD8+ but not CD4+ or DN MAIT cells are associated with improved overall survival

We next sought to determine if the improved clinical responses observed in patients with high frequencies of MAIT cells were associated with improved overall survival. To reduce the confounding factors of stage and treatment, only stage IV melanoma patients who received ≥ 3 doses of anti-PD1 and were on treatment at the time of the blood draw were included in this analysis (n = 27). Survival was calculated from the time of the blood draw and all blood draws occurred while the patients were still receiving therapy. The median follow-up time for this group of patients was 1.5 years (0.084–2.3 years). We found that patients with high levels of either total MAIT cells or CD8+ MAIT cells while on anti-PD1 therapy have significantly improved overall survival (OS) compared to those with low frequencies of these populations. In contrast, patients with high frequencies of circulating CD4+ MAIT cells have significantly decreased OS (Fig. 4f). These results demonstrate that the overall frequencies of different MAIT cell subsets in the circulation of melanoma patients undergoing anti-PD1 immunotherapy represent a potential novel prognostic correlate of immunotherapy success. While our study was conducted with only 27 stage IV patients, studies in a larger cohort of patients will be required to formally establish such findings. Furthermore, experiments aimed at defining the transcriptional differences between MAIT cell subsets in humans are currently underway and should reveal a potential explanation for our findings. Previous studies have not always separated MAIT cells into subsets based on CD4 and CD8 expression, which may partially explain some discrepancies regarding the role of MAIT cells in cancer (Duan et al., 2019; Yan et al., 2020). In our study, CD4+ MAIT cells showed decreased cytokine production (*p* = 0.0091), while cytokine production was unchanged in CD8+ MAIT cells in melanoma patients (Fig. 1f). These results suggest that CD8+ MAIT cells might be more resistant to melanoma-mediated suppression and potentially point to an antitumor role for these cells. Altogether, our study is the first to show differences in treatment response and survival of melanoma patients treated with anti-PD1 based upon the frequency of circulating MAIT cells. While previous studies of MAIT cells have correlated treatment responses with the frequencies of MAIT cells (Duan et al., 2019; Li et al., 2020), these previous studies did not focus on anti-PD1 therapy.

## Concluding remarks

The results of our study indicate that MAIT cells represent a positive predictive marker of clinical responses and overall survival in melanoma patients. While some aspects of these results diverge from the current literature in other cancers, the strikingly improved overall survival of patients with high levels of circulating CD8+ MAIT cells provides strong rationale for further investigation regarding the potential role of MAIT cells in antitumor immunity. Microbial exposure has been shown to impact the abundance of MAIT cells, and subsequent interactions with the microbiota can modulate their function (Constantinides et al., 2019; Legoux et al., 2019).

Although we did not examine the microbiome in this patient cohort, it is tempting to speculate that the correlations between the contents of the microbiome and responses to anti-PD1 immunotherapy (Gopalakrishnan et al., 2018; McQuade et al., 2019), might be, at least partly, mediated by microbial effects on MAIT cells. Nevertheless, this study should be interpreted considering its limitations, such as the relatively low numbers of matched samples from patients pre- and on-therapy. It is also important to note that melanoma tumors analyzed in the study were not responding to therapy at the time they were removed, and MAIT cells may be significantly different in tumors that respond to therapy. Additionally, this was a small single-center study meaning there may be inherent bias in a tertiary referral population. Despite these limitations, our results highlight the potential role of MAIT cells in antitumor immunity.

## Materials and methods

### Patient selection, ethics approval, and sample collection

Melanoma patients were identified at The University of Colorado Cancer Center. All patients provided written informed consent for sample and clinical data collection (Supplementary Table 1) according to the Colorado Multiple Institutional Review Board (COMIRB) Protocol #05–0309. Peripheral blood was obtained from healthy adult donors from the Children’s Hospital Colorado Blood Bank COMIRB #16–2367 and #17–0110. Peripheral blood was collected in acid citrate Vacutainer tubes (BD). Clinical responses to anti-PD1 therapy were measured according to RECIST 1.1 criteria by the treating oncologist using standard of care clinical imaging. Melanoma tumor samples were collected as part of standard of care surgical resections. Absolute numbers of different cells types were calculated from complete blood counts (CBC) performed as part of standard of care clinical visits on the same day as the research blood draws for this study.

### Sample preparation

PBMCs were isolated by Ficoll density centrifugation. Tumors were minced to produce pieces < 1mm^3^ and then incubated with 5µg/mL Liberase DL (Roche) at 37°C for 40 minutes and passed through 40μ cell strainer to create a single cell suspension.

### Flow cytometric analysis

MAIT cells were identified from whole blood by first incubating with the PE-conjugated 5-OP-RU-loaded MR1 tetramer (NIH Tetramer Core Facility) for 40 minutes at room temperature (Corbett et al., 2014). Samples were then stained with the Live/Dead discrimination dye Ghost Dye 780 (Tonbo Biosciences) and the fluorochrome-conjugated antibodies CD3-PerCP (SK7), CD4-Alexafluor 700 (OKT4), CD8-PE-Cy7 (SK1), CD25-BV605 (BC96), CD45-BV510 (HI30), and CD161-APC (HP-3G10), CD154-BV711 (24–31), PD1-BV421 (EH12.2H7), TCR Va7.2-FITC (3C10), and CD39-BV711 (TU66) in FACS buffer (PBS + 2% FBS) for 20 minutes at room temperature. Erythrocytes were lysed using BD Biosciences Pharm Lyse Buffer according to the manufacturer’s protocol.

For intracellular staining, PBMCs were incubated with phorbol 12-myristate 13-acetate (PMA, 50ng/mL Sigma Aldrich) and ionomycin (500ng/mL, Sigma Aldrich) in the presence of monensin (2μM, Biolegend) for 4 hours. Cells were washed twice in FACS buffer followed by staining with the MR1 tetramer and then cell surface staining with Live/Dead Ghost 780, CD3-PerCP (SK7), CD4-Alexafluor 700 (OKT4), CD8-PE-Cy7 (SK1), CD45-BV510 (HI30), and CD161-APC (HP-3G10), as described above. Cells were then fixed and permeabilized using the Intracellular Fixation and Permeabilization Buffer Set (eBioscience) according to the manufacturer’s protocol. Fixed and permeabilized cells were stained for GZMB-PE-Cy7 (QA16A02), GZMKFITC (GM26E7), Ki-67-Alexafluor 488 (Ki-67), IFNγ-BV421 (4S.B3), and TNFα-BV605 (Mab11), in permeabilization buffer (eBioscience) for 20 minutes at room temperature. All samples were analyzed on a Beckman Coulter CytoFlex S flow cytometer and data was analyzed using FlowJo Software (TreeStar).

### Statistical analysis

Statistical analyses were performed using one-way ANOVAs and two-tailed paired as well as unpaired *t*-tests in Prism (GraphPad). Error bars represent standard error of the mean (SEM). High and low levels of cell types were identified using the Evaluate Cutpoints R package (Ogluszka et al., 2019), with statistical significance of survival based on high or low levels determined using the Log-rank test in Prism (Graphpad). Levels of significance are denoted as follows: not significant (ns); *, *p* < 0.05; **, *p* < 0.01; ***, *p* < 0.001; and ****, *p* < 0.0001.

## Data Availability

All data referred to in the manuscript is included in the figures and supplementary information.

---

**Supplementary Figure 1.** Comparisons of circulating non-MAIT (NM)-CD8+ (a) or CD4+ (b) T cells in healthy donors (HD) and treatment naïve (TN) melanoma patients. Comparisons of the percentage of granzyme B (GZMB) (c) or granzyme K (GZMK) (d) in total MAIT cells or NM-CD8+ T cells. (e) Comparisons of the geometric mean fluorescence intensity (gMFI) of CD154 on circulating or tumor infiltrating MAIT cell populations. (f) Quantification of the absolute number of circulating double negative (DN) MAIT cells and NMCD8+ T cells. (g) Comparisons of the ratios of DN, CD4+, and CD8+ MAIT cells as a percentage of the total MAIT cell population.

**Supplementary Table 1.** Patient characteristics for blood samples.

## Acknowledgements

This work was supported by The National Institutes of Health R21CA245432 to MDM, CL, LG, and TMM. Further support was provided by the University of Colorado Anschutz Medical Campus Academic Enrichment Fund, University of Colorado GI and Liver Innate Immunity Program, the Colorado Clinical and Translation Sciences Institute, The Moore Family Foundation, the Patten-Davis Foundation, and University of Colorado Cancer Center Support Grant (P30CA046934).

